# Effectiveness of the BNT162b2 vaccine against SARS-CoV-2 infection among children and adolescents in Qatar

**DOI:** 10.1101/2022.07.26.22278045

**Authors:** Hiam Chemaitelly, Sawsan AlMukdad, Houssein H. Ayoub, Heba N. Altarawneh, Peter Coyle, Patrick Tang, Hadi M. Yassine, Hebah A. Al-Khatib, Maria K. Smatti, Mohammad R. Hasan, Zaina Al-Kanaani, Einas Al-Kuwari, Andrew Jeremijenko, Anvar Hassan Kaleeckal, Ali Nizar Latif, Riyazuddin Mohammad Shaik, Hanan F. Abdul-Rahim, Gheyath K. Nasrallah, Mohamed Ghaith Al-Kuwari, Hamad Eid Al-Romaihi, Adeel A. Butt, Mohamed H. Al-Thani, Abdullatif Al-Khal, Roberto Bertollini, Laith J. Abu-Raddad

**Author notes:** Correspondence to Dr. Hiam Chemaitelly, or Professor Laith J. Abu-Raddad,.

## Abstract

**Background:** The BNT162b2 COVID-19 vaccine is authorized for children 5-11 years of age and adolescents 12-17 years of age, but in different dose sizes. We assessed BNT162b2 real-world effectiveness against SARS-CoV-2 infection among children and adolescents in Qatar.

**Methods:** Three matched, retrospective, target-trial, cohort studies were conducted to compare incidence of SARS-CoV-2 infection in the national cohort of vaccinated individuals to incidence in the national cohort of unvaccinated individuals. Associations were estimated using Cox proportional-hazards regression models.

**Results:** Effectiveness of the 10 µg dose for children against Omicron infection was 25.7% (95% CI: 10.0-38.6%). It was highest at 49.6% (95% CI: 28.5-64.5%) right after the second dose, but waned rapidly thereafter and was negligible after 3 months. Effectiveness was 46.3% (95% CI: 21.5-63.3%) among those aged 5-7 years and 16.6% (−4.2-33.2%) among those aged 8-11 years. Effectiveness of the 30 µg dose for adolescents against Omicron infection was 30.6% (95% CI: 26.9-34.1%), but many adolescents were vaccinated months earlier. Effectiveness waned with time after the second dose. Effectiveness was 35.6% (95% CI: 31.2-39.6%) among those aged 12-14 years and 20.9% (13.8-27.4%) among those aged 15-17 years. Effectiveness of the 30 µg dose for adolescents against pre-Omicron infection was 87.6% (95% CI: 84.0-90.4%) and waned relatively slowly after the second dose.

**Conclusions:** Pediatric vaccination is associated with modest and rapidly waning protection against Omicron infection. Adolescent vaccination is associated with stronger and more durable protection, perhaps because of the larger dose size. Age at such young age appears to play a role in determining vaccine protection, with greater protection observed in younger than older children or adolescents.

## Introduction

The BNT162b2^1^ (Pfizer-BioNTech) mRNA Coronavirus Disease 2019 (COVID-19) vaccine has been authorized for use among adolescents 12-17 years of age and children 5-11 years of age, but in two formulations with different dose sizes, 30 µg versus 10 µg, respectively.^1,2^ Qatar launched mass COVID-19 immunization campaigns using these vaccines, first among adolescents in several phases starting in spring of 2021, and in February 2022 among children 5-11 years of age.

We assessed real-world effectiveness of the two-dose primary-series against severe acute respiratory syndrome coronavirus 2 (SARS-CoV-2) infection for the pediatric 10 µg BNT162b2 vaccine among children and the 30 µg BNT162b2 vaccine among adolescents. This was done in a national study in Qatar, a country of 2.8 million people^3^ that experienced five SARS-CoV-2 waves dominated sequentially by the original virus,^3^ Alpha,^4^ Beta,^5^ Omicron BA.1 and BA.2,^6^ and currently Omicron BA.4 and BA.5,^7^ in addition to a prolonged low-incidence phase dominated by Delta.^8^

## Methods

### Study population and data sources

This study was conducted in the resident population of Qatar. It analyzed the national, federated databases for COVID-19 laboratory testing, vaccination, hospitalization, and death, retrieved from the integrated, nationwide, digital-health information platform. Databases include all SARS-CoV-2-related data and associated demographic information, with no missing information, since pandemic onset, such as all polymerase chain reaction (PCR) tests, and starting from January 5, 2022 onward, rapid antigen tests conducted at healthcare facilities. Qatar has unusually diverse demographics in that 89% of the population are expatriates from over 150 countries.^3^ More detailed descriptions of Qatar’s population and of the SARS-CoV-2 national databases have been reported previously.^3,5,6,9,10^

### Study design and cohorts

Three matched, retrospective cohort studies that emulated randomized “target” trials^10,11^ were conducted to investigate BNT162b2 effectiveness in children aged 5-11 years and in adolescents aged 12-17 years ≥14 days after the second vaccine dose. In each study, we compared incidence of infection or of severe,^12^ critical,^12^ or fatal^13^ COVID-19 in the national cohort of infection-naïve individuals who completed the two-dose primary-series of the BNT162b2 vaccine (designated as the vaccinated cohort) to the national control cohort of individuals who were infection-naïve and unvaccinated (designated as the control cohort).

Documentation of infection in all cohorts was based on positive PCR or rapid antigen tests. Laboratory methods are in Supplementary Appendix Section S1. Classification of COVID-19 case severity (acute-care hospitalizations),^12^ criticality (intensive-care-unit hospitalizations),^12^ and fatality^13^ followed World Health Organization guidelines (Section S2).

### Cohort matching and follow-up

Vaccinated and infection-naïve individuals were exact-matched in a one-to-one ratio by sex, age, nationality, and comorbidity count (none, one, two or more comorbidities) to unvaccinated and infection-naïve individuals in the control cohort, to account for known differences in SARS-CoV-2 risk of exposure in Qatar.^3,14-17^ Matching by these factors was previously shown to provide adequate control of differences in risk of infection exposure in Qatar in studies of different epidemiologic designs and that included control groups to test for null effects.^9,18-21^ Matching was also done by calendar month of the second vaccine dose for the vaccinated cohort and of SARS-CoV-2-negative test for the control cohort. That is, individuals who received their second dose in a specific month were matched to unvaccinated individuals who had a record of a SARS-CoV-2-negative test in that same calendar month (Figures S1-S3). This was to ensure that individuals had active presence in Qatar at the same time. Matching was performed iteratively to ensure that individuals in the control cohort were alive, infection-free, and unvaccinated at the start of follow-up.

Each matched pair was followed from the calendar day when the vaccinated individual completed 14 days after the second dose. For exchangeability,^10,22^ both members of each matched pair were censored when the vaccinated individual received a third (booster) vaccine dose, or when a control individual received a first vaccine dose. Accordingly, individuals were followed up until the first of any of the following events: a documented SARS-CoV-2 infection (defined as the first PCR-positive or rapid-antigen-positive test after the start of follow-up, regardless of symptoms), or a third-dose (booster) vaccination for primary-series vaccinated individuals (with matched-pair censoring), or first-dose vaccination for controls (with matched-pair censoring), or death, or end of study censoring.

### Children Omicron 10 µg BNT162b2 Study

This study estimated effectiveness of the pediatric 10 µg BNT162b2 vaccine against infection with an Omicron subvariant in children aged 5-11 years, as children vaccination started only after onset of the Omicron wave in Qatar.^6,10,23^ Any child who received two doses of this vaccine between February 3, 2022 (earliest record for two-dose vaccination in children) and July 12, 2022 (end of study) was eligible for inclusion in the vaccinated cohort provided that the individual had no record of infection before the start of follow-up, on the 14^th^ day after the second dose. Any individual with a SARS-CoV-2-negative test during the study was eligible for inclusion in the control cohort, provided that the individual had no record of infection or vaccination before the start of follow-up.

### Adolescents Pre-Omicron 30 µg BNT162b2 Study

This study estimated effectiveness of the 30 µg BNT162b2 vaccine against infection with a pre-Omicron variant in adolescents aged 12-17 years. Any adolescent who received two doses of the BNT162b2 vaccine between February 1, 2021 (earliest record for two-dose vaccination in adolescents) and November 30, 2021 (end of study) was eligible for inclusion in the vaccinated cohort, provided that the individual had no record of infection before the start of follow-up. Follow-up was from the 14^th^ day after the second dose until November 30, 2021 (first evidence of Omicron in Qatar^6,10,23^ to ensure that all incidence was due to a pre-Omicron variant). Any individual with a SARS-CoV-2-negative test during the study was eligible for inclusion in the control cohort, provided that the individual had no record of infection or vaccination before the start of follow-up.

### Adolescents Omicron 30 µg BNT162b2 Study

This study estimated effectiveness of the 30 µg BNT162b2 vaccine against infection with an Omicron subvariant in adolescents aged 12-17 years. Any adolescent who received two doses of the BNT162b2 vaccine between February 1, 2021 and July 12, 2022 (end of study) was eligible for inclusion in the vaccinated cohort, provided that the individual had no record of infection or a third-dose vaccination before the start of follow-up. Follow-up was from December 19, 2021 (onset of the Omicron wave in Qatar),^6,10,23^ if second-dose vaccination occurred ≥14 days before this date (before the Omicron wave) and from the 14^th^ day after the second dose otherwise. Any individual with a SARS-CoV-2-negative test between February 1, 2021 and July 12, 2022 was eligible for inclusion in the control cohort, provided that the individual had no record of infection or vaccination before the start of follow-up.

### Statistical analysis

Eligible and matched cohorts were described using frequency distributions and measures of central tendency, and were compared using standardized mean differences (SMDs). An SMD of ≤0.1 indicated adequate matching.^24^ Cumulative incidence of infection (defined as the proportion of individuals at risk, whose primary endpoint during follow-up was a breakthrough infection for the vaccinated cohort, or an infection for the control cohort) was estimated using the Kaplan– Meier estimator method.^25^ Incidence rate of infection in each cohort, defined as the number of identified infections divided by the number of person-weeks contributed by all individuals in the cohort, was estimated, with a corresponding 95% confidence interval (CI) using a Poisson log-likelihood regression model with the Stata 17.0 *stptime* command.

Hazard ratios, comparing incidence of infection in the cohorts and corresponding 95% CIs, were calculated using Cox regression adjusted for the matching factors with the Stata 17.0 *stcox* command. Schoenfeld residuals and log-log plots for survival curves were used to test the proportional-hazards assumption. CIs were not adjusted for multiplicity; thus, they should not be used to infer definitive differences between groups. Interactions were not considered. Vaccine effectiveness was estimated as the 1-adjusted hazard ratio.

Subgroup analyses were conducted to investigate durability of vaccine protection. Adjusted hazard ratios were calculated, stratified by month since second dose or by sub-cohort of those vaccinated at different times. Subgroup analyses were also conducted to investigate differences in vaccine protection among those aged 5-7 years compared to those aged 8-11 years, and those aged 12-14 years compared to those aged 15-17 years. Sensitivity analyses adjusting effectiveness estimates for differences in testing frequency between cohorts were conducted. Statistical analyses were conducted using Stata/SE version 17.0 (Stata Corporation, College Station, TX, USA).

### Oversight

Hamad Medical Corporation and Weill Cornell Medicine-Qatar Institutional Review Boards approved this retrospective study with a waiver of informed consent. The study was reported following Strengthening the Reporting of Observational Studies in Epidemiology (STROBE) guidelines. The STROBE checklist is found in Table S1.

## Results

### Children and adolescent vaccination

Between February 3, 2022 and July 12, 2022, 27,244 children aged 5-11 years received two doses of the pediatric 10 µg BNT162b2 vaccine. Median date of the first dose was March 23, 2022, and median date of the second dose was April 14, 2022. Median time between the first and second doses was 21 days (interquartile range (IQR), 21-21 days). None of the children received a third vaccine dose.

Between February 1, 2021 and July 12, 2022, 104,020 adolescents aged 12-17 years received at least two vaccine doses, of whom 102,536 (98.6%) received two doses of the 30 µg BNT162b2 vaccine. Median date of the first dose was June 28, 2021, and median date of the second dose was July 25, 2021. A total of 29,856 adolescents received a third (booster) dose. Median date of the third dose was March 18, 2022. Median time between the first and second doses was 28 days (IQR, 21-28 days) and between the second and third doses was 243 days (IQR, 215-284 days).

### Children Omicron 10 µg BNT162b2 Study

Figure S1 shows the study population selection process and Table 1 describes the full and matched cohort characteristics. Matched cohorts each included 18,728 children. The study was conducted on the total children population of Qatar; thus, the study population is broadly representative of the internationally diverse pediatric population of Qatar (Table S2).

**Table 1.** **Baseline characteristics of eligible and matched vaccinated and control cohorts in the study of effectiveness of the pediatric 10 µg BNT162b2 vaccine among children aged 5-11 years against infection with an Omicron subvariant (Children Omicron 10 µg BNT162b2 Study).**

| Characteristics | Full eligible cohorts |  |  | Matched cohorts* |  |  |
| --- | --- | --- | --- | --- | --- | --- |
|  | Vaccinated cohort | Control cohort | SMD† | Vaccinated cohort | Control cohort | SMD† |
|  | N=21,888 | N=91,870 |  | N=18,728 | N=18,728 |  |
| Median age (IQR)—years | 8 (7-10) | 8 (6-9) | 0.32‡ | 8 (7-10) | 8 (7-10) | 0.00‡ |
| Age—years |  |  |  |  |  |  |
| 5 | 1,874 (8.6) | 17,503 (19.1) |  | 1,802 (9.6) | 1,802 (9.6) |  |
| 6 | 2,768 (12.7) | 14,190 (15.5) |  | 2,512 (13.4) | 2,512 (13.4) |  |
| 7 | 3,175 (14.5) | 13,526 (14.7) |  | 2,769 (14.8) | 2,769 (14.8) |  |
| 8 | 3,381 (15.5) | 12,555 (13.7) | 0.36 | 2,850 (15.2) | 2,850 (15.2) | 0.00 |
| 9 | 3,331 (15.2) | 12,099 (13.2) |  | 2,847 (15.2) | 2,847 (15.2) |  |
| 10 | 3,593 (16.4) | 10,903 (11.9) |  | 2,946 (15.7) | 2,946 (15.7) |  |
| 11 | 3,766 (17.2) | 11,094 (12.1) |  | 3,002 (16.0) | 3,002 (16.0) |  |
| Sex |  |  |  |  |  |  |
| Male | 11,026 (50.4) | 46,950 (51.1) | 0.01 | 9,374 (50.1) | 9,374 (50.1) | 0.00 |
| Female | 10,862 (49.6) | 44,920 (48.9) |  | 9,354 (49.9) | 9,354 (49.9) |  |
| Nationality§ |  |  |  |  |  |  |
| Bangladeshi | 567 (2.6) | 1,033 (1.1) |  | 373 (2.0) | 373 (2.0) |  |
| Egyptian | 718 (3.3) | 10,289 (11.2) |  | 718 (3.8) | 718 (3.8) |  |
| Filipino | 2,918 (13.3) | 2,123 (2.3) |  | 1,542 (8.2) | 1,542 (8.2) |  |
| Indian | 12,822 (58.6) | 23,924 (26.0) |  | 11,951 (63.8) | 11,951 (63.8) |  |
| Nepalese | 144 (0.7) | 193 (0.2) | 1.18 | 76 (0.4) | 76 (0.4) | 0.00 |
| Pakistani | 1,227 (5.6) | 5,607 (6.1) |  | 1,190 (6.4) | 1,190 (6.4) |  |
| Qatari | 128 (0.6) | 18,921 (20.6) |  | 128 (0.7) | 128 (0.7) |  |
| Sri Lankan | 350 (1.6) | 1,096 (1.2) |  | 311 (1.7) | 311 (1.7) |  |
| Sudanese | 95 (0.4) | 2,837 (3.1) |  | 95 (0.5) | 95 (0.5) |  |
| Other nationalities¶ | 2,919 (13.3) | 25,847 (28.1) |  | 2,344 (12.5) | 2,344 (12.5) |  |
| Comorbidity count |  |  |  |  |  |  |
| None | 17,954 (82.0) | 71,929 (78.3) |  | 16,030 (85.6) | 16,030 (85.6) |  |
| 1 | 3,345 (15.3) | 15,973 (17.4) | 0.11 | 2,362 (12.6) | 2,362 (12.6) | 0.00 |
| 2+ | 589 (2.7) | 3,968 (4.3) |  | 336 (1.8) | 336 (1.8) |  |

IQR denotes interquartile range, SARS-CoV-2 severe acute respiratory syndrome coronavirus 2, and SMD standardized mean difference.
\*Vaccinated and control cohorts were exact-matched in a 1:1 ratio by sex, age, nationality, comorbidity count, and calendar month of the second vaccine dose/SARS-CoV-2 test.
†SMD is the difference in the mean of a covariate between groups divided by the pooled standard deviation. An SMD ≤0.1 indicates adequate matching.
‡SMD is for the mean difference between groups divided by the pooled standard deviation.
§Nationalities were chosen to represent the most populous groups in Qatar.
¶These comprise up to 140 other nationalities in the unmatched cohorts, and 66 other nationalities in the matched cohorts.

Median time of follow-up was 69 days (IQR, 31-97 days) for the vaccinated cohort and 69 days (IQR, 30-97 days) for the control cohort (Figure 1). There were 184 infections in the vaccinated cohort and 248 infections in the control cohort during follow-up (Figure S1). None of these infections progressed to severe, critical, or fatal COVID-19. Infection incidence coincided with time when incidence was due to Omicron BA.1, BA.2, BA.4, or BA.5 subvariants, but after the peak of the major BA.1 and BA.2 wave in mid-January of 2022.^6,7,10,23^ Cumulative incidence of infection was estimated at 2.1% (95% CI: 1.7-2.4%) for the vaccinated cohort and 2.4% (95% CI: 2.0-2.7%) for the control cohort, 110 days after the start of follow-up (Figure 1A).

**Figure 1.**
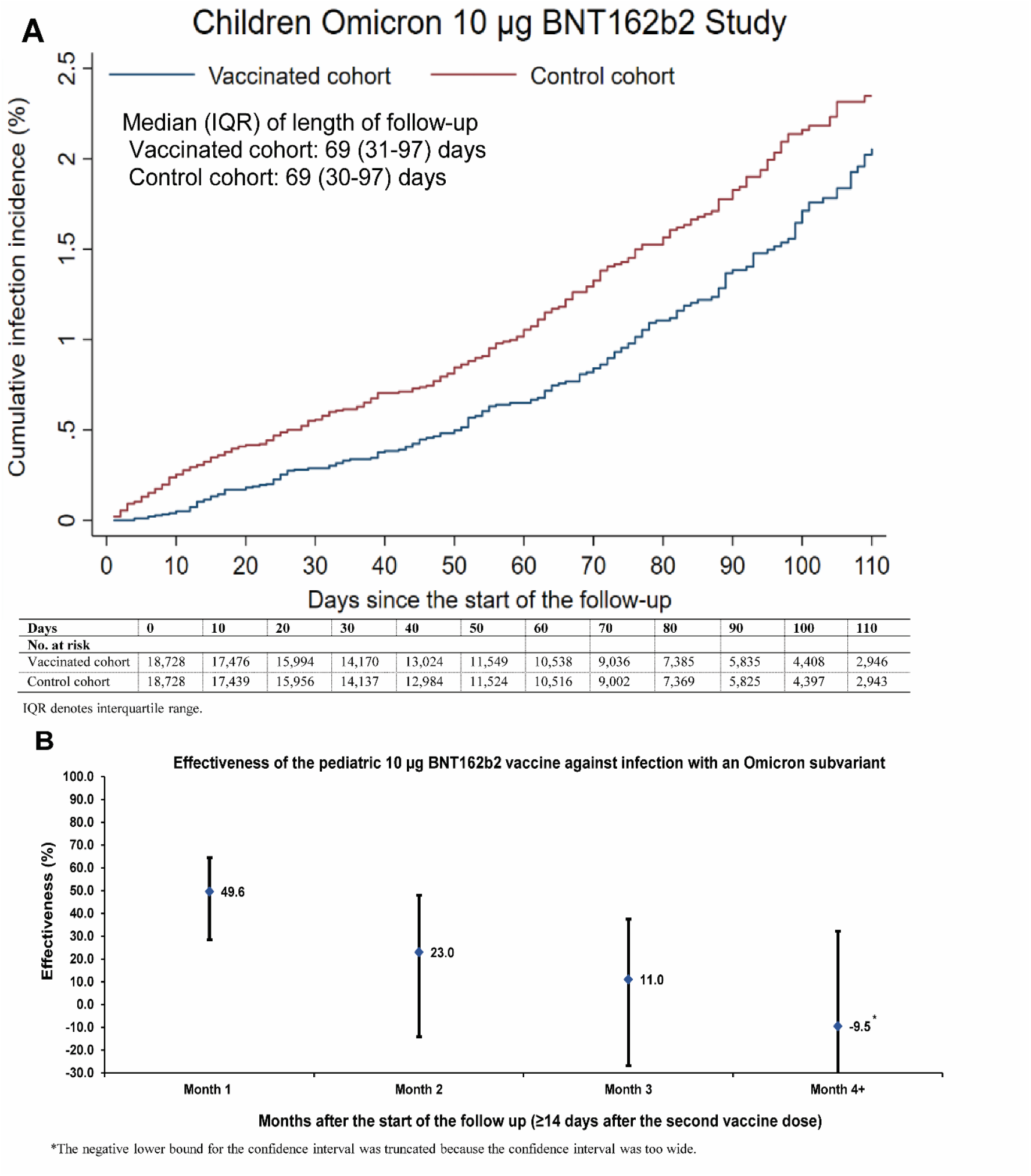
**A) Cumulative incidence of SARS-CoV-2 Omicron infection in children aged 5-11 years who received two doses of the pediatric 10 µg BNT162b2 vaccine compared to unvaccinated controls. B) Effectiveness against Omicron infection of the pediatric 10 µg BNT162b2 vaccine by month after second dose.**

The overall hazard ratio for infection, adjusted for sex, age, 10 nationality groups, comorbidity count, and calendar month of the second dose/SARS-CoV-2-negative test, was estimated at 0.74 (95% CI: 0.61-0.90; Table 2). Effectiveness of the pediatric 10 µg BNT162b2 vaccine against Omicron infection was estimated at 25.7% (95% CI: 10.0-38.6%). Effectiveness declined with time after second dose (Figure 1B). It was highest at 49.6% (95% CI: 28.5-64.5%) right after the second dose, but waned rapidly thereafter and was negligible after 3 months.

**Table 2.** BNT162b2 effectiveness against infection among children and adolescents in Qatar.

| **Epidemiological measures** | **Estimate**<br>**(95% CI)** | **Effectiveness in %**<br>**(95% CI)** |
| --- | --- | --- |
| **Children aged 5-11 years—Children Omicron 10 µg BNT162b2 Study** |  |  |
| Total follow-up time—Vaccinated cohort (person-weeks) | 173,451 | -- |
| Total follow-up time—Control cohort (person-weeks) | 173,082 | -- |
| Incidence rate of infection—Vaccinated cohort (per 10,000 person-weeks) | 10.6 (9.2 to 12.3) | -- |
| Incidence rate of infection—Control cohort (per 10,000 person-weeks) | 14.3 (12.7 to 16.2) | -- |
| Unadjusted hazard ratio for infection with an Omicron subvariant | 0.74 (0.61 to 0.90) | 25.7 (10.0 to 38.6) |
| Adjusted hazard ratio for infection with an Omicron subvariant* | 0.74 (0.61 to 0.90) | 25.7 (10.0 to 38.6) |
| **Adolescents aged 12-17 years—Adolescents Pre-Omicron 30 µg BNT162b2 Study** |  |  |
| Total follow-up time—Vaccinated cohort (person-weeks) | 192,309 | -- |
| Total follow-up time—Control cohort (person-weeks) | 185,751 | -- |
| Incidence rate of infection—Vaccinated cohort (per 10,000 person-weeks) | 3.5 (2.7 to 4.4) | -- |
| Incidence rate of infection—Control cohort (per 10,000 person-weeks) | 28.2 (25.8 to 30.7) | -- |
| Unadjusted hazard ratio for infection with a pre-Omicron variant | 0.13 (0.10 to 0.16) | 87.5 (83.8 to 90.3) |
| Adjusted hazard ratio for infection with a pre-Omicron variant* | 0.12 (0.10 to 0.16) | 87.6 (84.0 to 90.4) |
| **Adolescents aged 12-17 years—Adolescents Omicron 30 µg BNT162b2 Study** |  |  |
| Total follow-up time—Vaccinated cohort (person-weeks) | 338,838 | -- |
| Total follow-up time—Control cohort (person-weeks) | 319,291 | -- |
| Incidence rate of infection—Vaccinated cohort (per 10,000 person-weeks) | 74.4 (71.5 to 77.3) | -- |
| Incidence rate of infection—Control cohort (per 10,000 person-weeks) | 104.5 (101.0 to 108.1) | -- |
| Unadjusted hazard ratio for infection with an Omicron subvariant | 0.72 (0.69 to 0.76) | 27.7 (23.8 to 31.3) |
| Adjusted hazard ratio for infection with an Omicron subvariant* | 0.69 (0.66 to 0.73) | 30.6 (26.9 to 34.1) |

CI denotes confidence interval and SARS-CoV-2 severe acute respiratory syndrome coronavirus 2.
\*Cox regression analysis adjusted for sex, age, 10 nationality groups (Tables 1 and 3), comorbidity count (Tables 1 and 3), and calendar month of the second vaccine dose/SARS-CoV-2 test.

Stratified analyses by age group estimated the overall adjusted hazard ratio at 0.54 (95% CI: 0.37-0.79) among those aged 5-7 years and at 0.83 (95% CI: 0.67-1.04) among those aged 8-11 years. Corresponding vaccine effectiveness was 46.3% (95% CI: 21.5-63.3%) and 16.6% (−4.2-33.2%), respectively.

### Adolescents Pre-Omicron 30 µg BNT162b2 Study

Figure S2 shows the study population selection process and Table 3 describes the full and matched cohort characteristics. Matched cohorts each included 23,317 adolescents. The study was conducted on the total adolescent population of Qatar; thus, the study population is representative of Qatar’s adolescents (Table S2).

**Table 3.** **Baseline characteristics of the eligible and matched vaccinated and control cohorts in the study of effectiveness of the 30 µg BNT162b2 vaccine among adolescents aged 12-17 years against infection with a pre-Omicron variant (Adolescents Pre-Omicron 30 µg BNT162b2 Study) and against infection with an Omicron subvariant (Adolescents Omicron 30 µg BNT162b2 Study).**

| Characteristics | Pre-Omicron analysis (Adolescents Pre-Omicron 30 µg BNT162b2 Study) |  |  |  |  |  | Omicron analysis (Adolescents Omicron 30 µg BNT162b2 Study) |  |  |  |  |  |
| --- | --- | --- | --- | --- | --- | --- | --- | --- | --- | --- | --- | --- |
|  | Full eligible cohorts |  |  | Matched cohorts* |  |  | Full eligible cohorts |  |  | Matched cohorts* |  |  |
|  | Vaccinated cohort | Control cohort | SMD† | Vaccinated cohort | Control cohort | SMD† | Vaccinated cohort | Control cohort | SMD† | Vaccinated cohort | Control cohort | SMD† |
|  | N=81,647 | N=66,006 |  | N=23,317 | N=23,317 |  | N=89,505 | N=86,280 |  | N=17,903 | N=17,903 |  |
| Median age (IQR)—years | 15 (14-16) | 14 (12-15) | 0.50‡ | 14 (13-16) | 14 (13-16) | 0.00‡ | 15 (13-16) | 14 (12-15) | 0.47‡ | 13 (12-15) | 13 (12-15) | 0.00‡ |
| Age—years |  |  |  |  |  |  |  |  |  |  |  |  |
| 12 | 3,174 (3.9) | 18,636 (28.2) | 0.71 | 2,767 (11.9) | 2,767 (11.9) | 0.00 | 7,865 (8.8) | 28,852 (33.4) | 0.64 | 6,003 (33.5) | 6,003 (33.5) | 0.00 |
| 13 | 16,581 (20.3) | 11,190 (17.0) |  | 5,767 (24.7) | 5,767 (24.7) |  | 17,713 (19.8) | 13,530 (15.7) |  | 3,389 (18.9) | 3,389 (18.9) |  |
| 14 | 16,427 (20.1) | 10,382 (15.7) |  | 4,535 (19.5) | 4,535 (19.5) |  | 17,218 (19.2) | 12,403 (14.4) |  | 2,677 (15.0) | 2,677 (15.0) |  |
| 15 | 15,675 (19.2) | 9,861 (14.9) |  | 3,870 (16.6) | 3,870 (16.6) |  | 16,259 (18.2) | 11,866 (13.8) |  | 2,254 (12.6) | 2,254 (12.6) |  |
| 16 | 14,998 (18.4) | 8,257 (12.5) |  | 2,734 (11.7) | 2,734 (11.7) |  | 15,447 (17.3) | 10,136 (11.8) |  | 1,790 (10.0) | 1,790 (10.0) |  |
| 17 | 14,792 (18.1) | 7,680 (11.6) |  | 3,644 (15.6) | 3,644 (15.6) |  | 15,003 (16.8) | 9,493 (11.0) |  | 1,790 (10.0) | 1,790 (10.0) |  |
| Sex |  |  |  |  |  |  |  |  |  |  |  |  |
| Male | 41,919 (51.3) | 34,594 (52.4) | 0.02 | 12,083 (51.8) | 12,083 (51.8) | 0.00 | 45,936 (51.3) | 44,941 (52.1) | 0.02 | 9,258 (51.7) | 9,258 (51.7) | 0.00 |
| Female | 39,728 (48.7) | 31,412 (47.6) |  | 11,234 (48.2) | 11,234 (48.2) |  | 43,569 (48.7) | 41,339 (47.9) |  | 8,645 (48.3) | 8,645 (48.3) |  |
| Nationality§ |  |  |  |  |  |  |  |  |  |  |  |  |
| Bangladeshi | 1,054 (1.3) | 967 (1.5) | 0.26 | 391 (1.7) | 391 (1.7) | 0.00 | 1,182 (1.3) | 1,521 (1.8) | 0.24 | 414 (2.3) | 414 (2.3) | 0.00 |
| Egyptian | 8,894 (10.9) | 7,538 (11.4) |  | 3,379 (14.5) | 3,379 (14.5) |  | 9,835 (11.0) | 9,460 (11.0) |  | 2,051 (11.5) | 2,051 (11.5) |  |
| Filipino | 3,029 (3.7) | 1,118 (1.7) |  | 280 (1.2) | 280 (1.2) |  | 3,372 (3.8) | 1,896 (2.2) |  | 345 (1.9) | 345 (1.9) |  |
| Indian | 12,943 (15.9) | 6,569 (10.0) |  | 2,812 (12.1) | 2,812 (12.1) |  | 15,084 (16.9) | 9,434 (10.9) |  | 2,735 (15.3) | 2,735 (15.3) |  |
| Nepalese | 102 (0.1) | 46 (0.1) |  | 9 (0.04) | 9 (0.04) |  | 120 (0.1) | 76 (0.1) |  | 7 (0.04) | 7 (0.04) |  |
| Pakistani | 5,089 (6.2) | 2,876 (4.4) |  | 982 (4.2) | 982 (4.2) |  | 5,622 (6.3) | 3,932 (4.6) |  | 1,051 (5.9) | 1,051 (5.9) |  |
| Qatari | 22,927 (28.1) | 22,196 (33.6) |  | 7,531 (32.3) | 7,531 (32.3) |  | 24,210 (27.1) | 26,578 (30.8) |  | 5,024 (28.1) | 5,024 (28.1) |  |
| Sri Lankan | 603 (0.7) | 260 (0.4) |  | 73 (0.31) | 73 (0.31) |  | 700 (0.8) | 494 (0.6) |  | 100 (0.6) | 100 (0.6) |  |
| Sudanese | 3,130 (3.8) | 2,253 (3.4) |  | 829 (3.6) | 829 (3.6) |  | 3,395 (3.8) | 2,855 (3.3) |  | 616 (3.4) | 616 (3.4) |  |
| Other nationalities¶ | 23,876 (29.2) | 22,183 (33.6) |  | 7,031 (30.2) | 7,031 (30.2) |  | 25,985 (29.0) | 30,034 (34.8) |  | 5,560 (31.1) | 5,560 (31.1) |  |
| Comorbidity count |  |  |  |  |  |  |  |  |  |  |  |  |
| None | 64,614 (79.1) | 52,741 (79.9) | 0.02 | 19,183 (82.3) | 19,183 (82.3) | 0.00 | 71,144 (79.5) | 70,646 (81.9) | 0.06 | 15,185 (84.8) | 15,185 (84.8) | 0.00 |
| 1-2 | 13,478 (16.5) | 10,344 (15.7) |  | 3,309 (14.2) | 3,309 (14.2) |  | 14,592 (16.3) | 12,239 (14.2) |  | 2,208 (12.3) | 2,208 (12.3) |  |
| 3+ | 3,555 (4.4) | 2,921 (4.4) |  | 825 (3.5) | 825 (3.5) |  | 3,769 (4.2) | 3,395 (3.9) |  | 510 (2.9) | 510 (2.9) |  |

IQR denotes interquartile range, SARS-CoV-2 severe acute respiratory syndrome coronavirus 2, and SMD standardized mean difference.
\*Vaccinated and control cohorts were exact-matched in a 1:1 ratio by sex, age, nationality, comorbidity count, and calendar month of the second vaccine dose/SARS-CoV-2 test.
†SMD is the difference in the mean of a covariate between groups divided by the pooled standard deviation. An SMD ≤0.1 indicates adequate matching.
‡SMD is for the mean difference between groups divided by the pooled standard deviation.
§Nationalities were chosen to represent the most populous groups in Qatar.
¶These comprise up to 135 other nationalities in the unmatched cohorts, and 75 other nationalities in the matched cohorts in the pre-Omicron analysis. These also comprise up to 143 other nationalities in the unmatched cohorts, and 73 other nationalities in the matched cohorts in the Omicron analysis.

Median time of follow-up was 45 days (IQR, 16-88 days) for the vaccinated cohort and 43 days (IQR, 15-85 days) for the control cohort (Figure 2A). There were 67 infections in the vaccinated cohort and 523 infections in the control cohort during follow-up (Figure S2). None of these infections progressed to severe, critical, or fatal COVID-19. Infection incidence coincided with time when Alpha, Beta, and especially Delta dominated incidence.^8,9,26,27^ Cumulative incidence of infection was estimated at 0.8% (95% CI: 0.6-1.0%) for the vaccinated cohort and at 4.1% (95% CI: 3.7-4.6%) for the control cohort, 135 days after the start of follow-up (Figure 2A).

**Figure 2.**
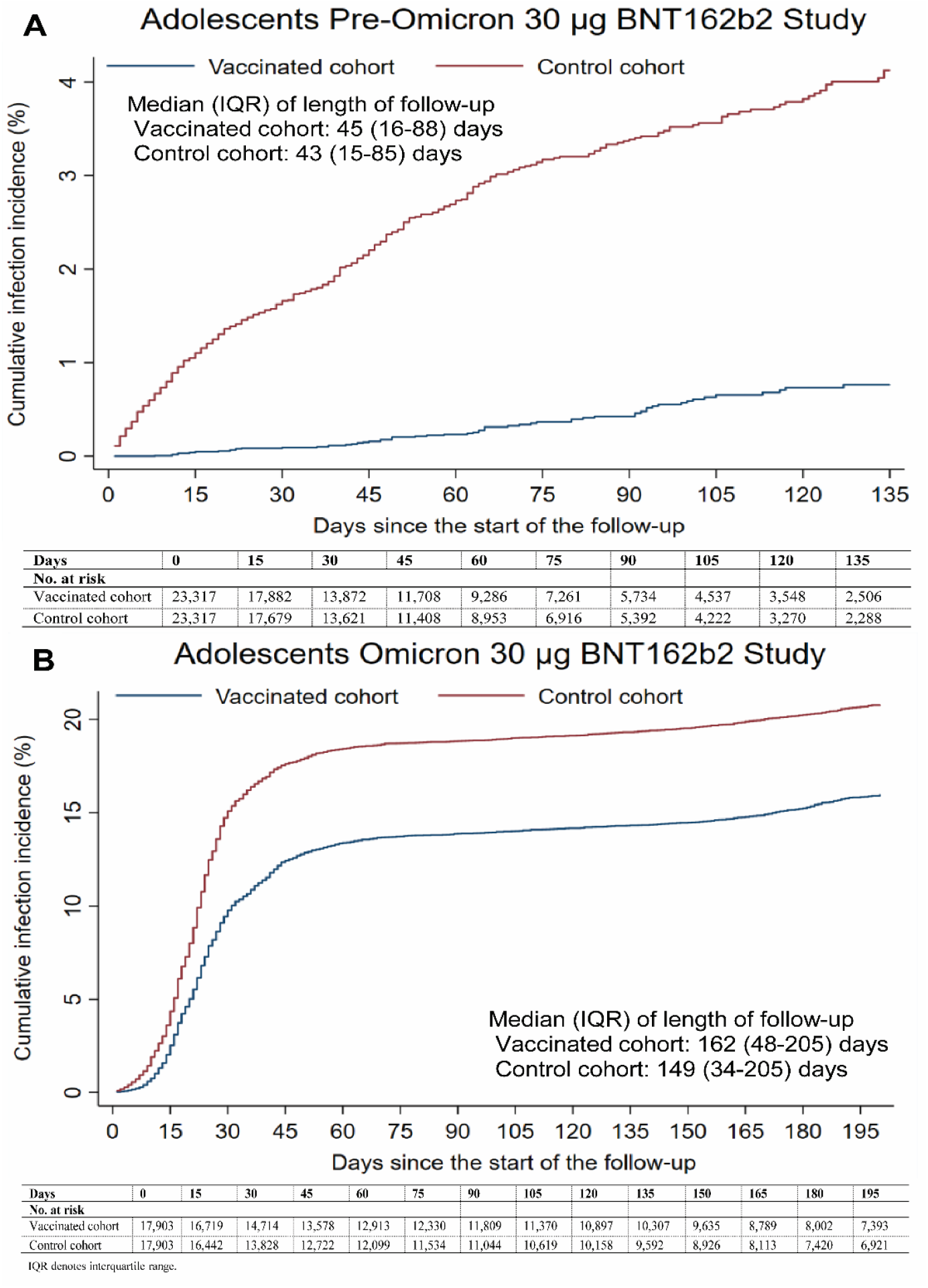
**Cumulative incidence of SARS-CoV-2 infection in adolescents aged 12-17 years who received two doses of the 30 µg BNT162b2 vaccine compared to unvaccinated controls in the A) pre-Omicron study (Adolescents Pre-Omicron 30 µg BNT162b2 Study) and B) Omicron study (Adolescents Omicron 30 µg BNT162b2 Study).**

The overall adjusted hazard ratio for infection was estimated at 0.12 (95% CI: 0.10-0.16; Table 2). Effectiveness of the 30 µg BNT162b2 vaccine against infection with a pre-Omicron variant was estimated at 87.6% (95% CI: 84.0-90.4%). Effectiveness declined with time after the second dose (Figure 3A). It was highest at 95.3% (95% CI: 92.0-97.2%) right after the second dose, but waned (relatively) slowly thereafter.

**Figure 3.**
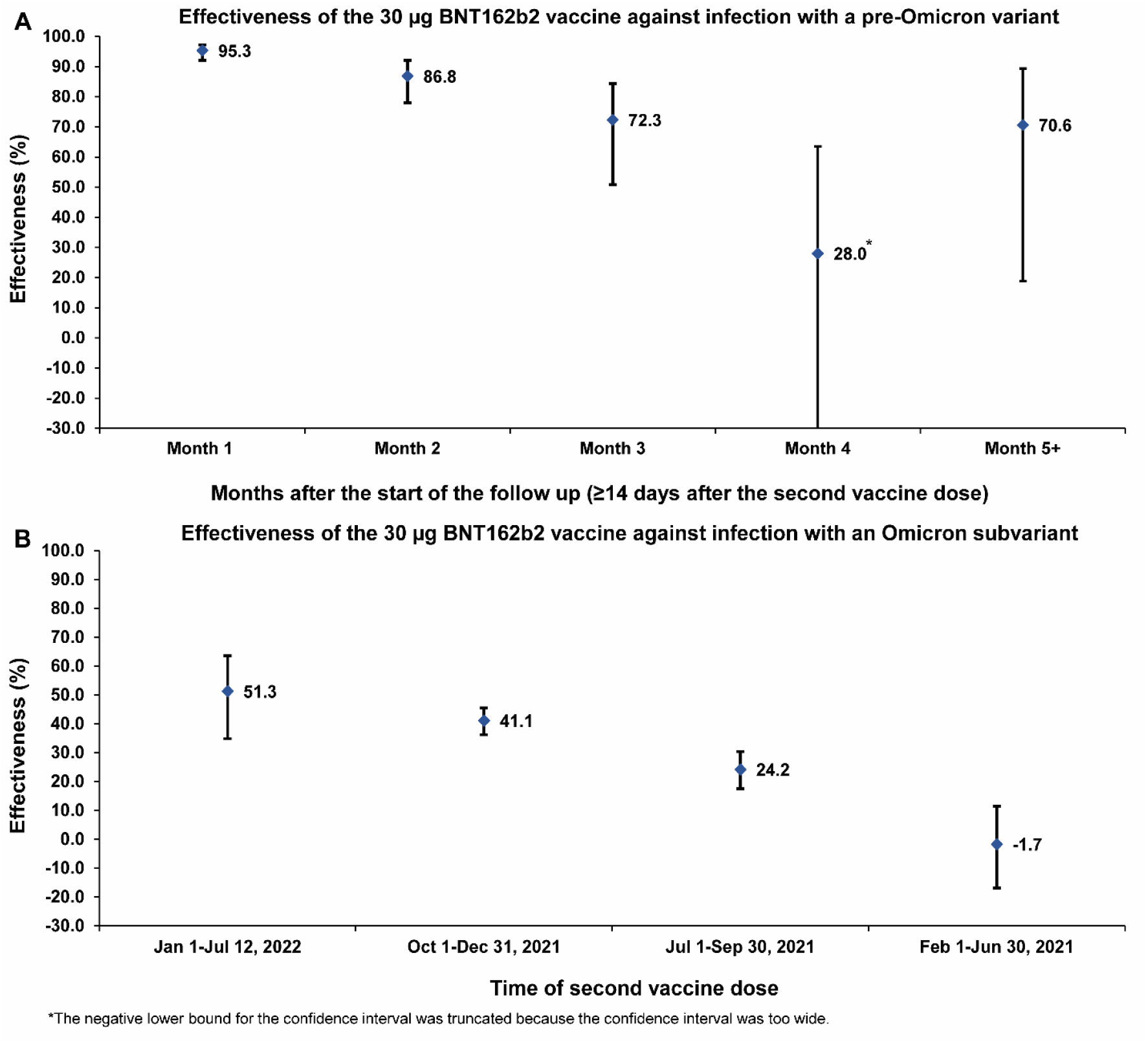
**A) Effectiveness against pre-Omicron infection of the 30 µg BNT162b2 vaccine for adolescents by month after the second dose. B) Effectiveness against Omicron infection of the 30 µg BNT162b2 vaccine for adolescents by sub-cohort of those vaccinated at different times.**

Stratified analyses by age group estimated the overall adjusted hazard ratio at 0.11 (95% CI: 0.07-0.16) among those aged 12-14 years and at 0.14 (95% CI: 0.10-0.20) among those aged 15-17 years. Corresponding vaccine effectiveness was 89.4% (95% CI: 84.5-92.7%) and 85.7% (79.7-89.8%), respectively.

### Adolescents Omicron 30 µg BNT162b2 Study

Figure S3 shows the study population selection process and Table 3 describes the full and matched cohort characteristics. Matched cohorts each included 17,903 adolescents. The study is representative of the adolescent population of Qatar (Table S2).

Median time of follow-up was 162 days (IQR, 48-205 days) for the vaccinated cohort and 149 days (IQR, 34-205 days) for the control cohort (Figure 2B). There were 2,520 infections in the vaccinated cohort during follow-up, of which one progressed to critical COVID-19, and 3,337 infections in the control cohort, of which one progressed to severe and another to critical

COVID-19 (Figure S3). Infection incidence coincided initially with the major Omicron BA.1 and BA.2 wave in January of 2022 (Figure 2B).^6,10,23^ Subsequently, infection incidence was due to BA.1, BA.2, BA.4, or BA.5 subvariants.^6,7,10,23^ Cumulative incidence of infection was estimated at 15.9% (95% CI: 15.3-16.4%) for the vaccinated cohort and at 20.7% (95% CI: 20.1-21.3%) for the control cohort, 195 days after the start of follow-up (Figure 2B).

The overall adjusted hazard ratio for infection was estimated at 0.69 (95% CI: 0.66-0.73; Table 2). Effectiveness of the 30 µg BNT162b2 vaccine against infection with an Omicron subvariant was estimated at 30.6% (95% CI: 26.9-34.1%). Effectiveness declined with time after the second dose (Figure 3B). It was highest at 51.3% (95% CI: 34.9-63.6%) for those who had their second dose recently, but negligible for those who completed their primary series between February 1, 2021 and June 30, 2021.

Stratified analyses by age group estimated the overall adjusted hazard ratio at 0.64 (95% CI: 0.60-0.69) among those aged 12-14 years and at 0.79 (95% CI: 0.73-0.86) among those aged 15-17 years. Corresponding vaccine effectiveness was 35.6% (95% CI: 31.2-39.6%) and 20.9% (13.8-27.4%), respectively.

## Discussion

The pediatric 10 µg BNT162b2 dose for children was associated with only modest protection against infection with an Omicron subvariant at ∼25%. This protection was also short lived, declining from ∼50% right after the second dose to negligible levels after 3 months. Meanwhile, the 30 µg BNT162b2 dose for adolescents was associated with stronger protection against Omicron infection and slower waning, suggesting a critical role of vaccine dose size. Overall protection of the 30 µg BNT162b2 dose was ∼30%, but many adolescents were vaccinated months earlier. For adolescents vaccinated concurrently with children, protection was at ∼50%, twice that for children.

Protection of the 30 µg BNT162b2 dose was stronger against infection with a pre-Omicron variant and waned (relatively) slowly. Protection was at ∼95% right after the second dose and remained strong at >50% for at least 5 months. Overall, protection and waning patterns of the 30 µg BNT162b2 dose among adolescents paralleled those among adults, though protection was slightly stronger among adolescents.^9,28^

Age appeared to play a role in determining the level of protection. Across analyses, vaccine protection against Omicron infection was higher in younger than older children or adolescents. For children, protection was at ∼45% among those aged 5-7 years, but only ∼15% among those aged 8-11 years. For adolescents, protection was at ∼35% among those aged 12-14 years, but only ∼20% among those aged 15-17 years. These findings are consistent with evidence on vaccine protection among children and adolescents in other countries.^2,29-33^

This study has limitations. With the lower severity of SARS-CoV-2 infection among children^32,34^ and the lower severity of Omicron infections,^35,36^ there were too few severe,^12^ critical,^12^ or fatal^13^ COVID-19 cases to estimate vaccine effectiveness against severe forms of COVID-19. We investigated incidence of documented infections, but other infections may have occurred and gone undocumented. Undocumented infections confer immunity or boost existing immunity, thereby perhaps affecting estimates of vaccine effectiveness, if differing among cohorts.^37^ Testing frequency differed between cohorts, mainly related to different testing guidelines for travel for vaccinated and unvaccinated individuals, but sensitivity analyses adjusting for these differences showed overall similar findings (Table S3).

As an observational study, investigated cohorts were neither blinded nor randomized, so unmeasured or uncontrolled confounding cannot be excluded. While cohorts were matched by sex, age, nationality, and comorbidity count, this was not possible for other factors such as geography, as such data were unavailable. However, Qatar is essentially a city state and infection incidence was broadly distributed across neighborhoods.

Matching was done to control for factors that affect infection exposure in Qatar.^3,14-17^ The matching prescription had already been investigated in previous studies of different epidemiologic designs, and using control groups to test for null effects.^9,18-21^ These control groups included unvaccinated cohorts versus vaccinated cohorts within two weeks of the first dose,^9,18-20^ when vaccine protection is negligible,^1^ and mRNA-1273-versus BNT162b2-vaccinated cohorts, also in the first two weeks after the first dose.^21^ These studies have shown that this prescription provides adequate control of differences in infection exposure.^9,18-21^ The above analyses were implemented using Qatar’s total population of children and adolescents with large sample sizes; thus, perhaps minimizing the likelihood of bias.

In conclusion, the pediatric 10 µg BNT162b2 dose for children is associated with modest and rapidly waning protection against Omicron infection. Meanwhile, the 30 µg BNT162b2 dose for adolescents is associated with a stronger and more durable protection, suggesting a critical role of dose size in determining vaccine protection. Protection of the 30 µg BNT162b2 dose was strong against infection with a pre-Omicron variant and waned relatively slowly. Age at such young age appears to influence vaccine protection, with higher protection observed in younger than older children or adolescents.

### Sources of support and acknowledgements

We acknowledge the many dedicated individuals at Hamad Medical Corporation, the Ministry of Public Health, the Primary Health Care Corporation, Qatar Biobank, Sidra Medicine, and Weill Cornell Medicine-Qatar for their diligent efforts and contributions to make this study possible. The authors are grateful for institutional salary support from the Biomedical Research Program and the Biostatistics, Epidemiology, and Biomathematics Research Core, both at Weill Cornell Medicine-Qatar, as well as for institutional salary support provided by the Ministry of Public Health, Hamad Medical Corporation, and Sidra Medicine. The authors are also grateful for the Qatar Genome Programme and Qatar University Biomedical Research Center for institutional support for the reagents needed for the viral genome sequencing. The funders of the study had no role in study design, data collection, data analysis, data interpretation, or writing of the article. Statements made herein are solely the responsibility of the authors.

## Data Availability

The dataset of this study is a property of the Qatar Ministry of Public Health that was provided to the researchers through a restricted-access agreement that prevents sharing the dataset with a third party or publicly. Future access to this dataset can be considered through a direct application for data access to Her Excellency the Minister of Public Health (https://www.moph.gov.qa/english/Pages/default.aspx). Aggregate data are available within the manuscript and its Supplementary information.

## Author contributions

HC co-designed the study, performed the statistical analyses, and co-wrote the first draft of the article. LJA conceived and co-designed the study, led the statistical analyses, and co-wrote the first draft of the article. PT and MRH conducted multiplex, RT-qPCR variant screening and viral genome sequencing. HY, HAK, and MS conducted viral genome sequencing. All authors contributed to data collection and acquisition, database development, discussion and interpretation of the results, and to the writing of the manuscript. All authors have read and approved the final manuscript.

## Supplementary Appendix

## Section S1. Laboratory methods and variant ascertainment

### Real-time reverse-transcription polymerase chain reaction testing

Nasopharyngeal and/or oropharyngeal swabs were collected for polymerase chain reaction (PCR) testing and placed in Universal Transport Medium (UTM). Aliquots of UTM were: 1) extracted on KingFisher Flex (Thermo Fisher Scientific, USA), MGISP-960 (MGI, China), or ExiPrep 96 Lite (Bioneer, South Korea) followed by testing with real-time reverse-transcription PCR (RT-qPCR) using TaqPath COVID-19 Combo Kits (Thermo Fisher Scientific, USA) on an ABI 7500 FAST (Thermo Fisher Scientific, USA); 2) tested directly on the Cepheid GeneXpert system using the Xpert Xpress SARS-CoV-2 (Cepheid, USA); or 3) loaded directly into a Roche cobas 6800 system and assayed with the cobas SARS-CoV-2 Test (Roche, Switzerland). The first assay targets the viral S, N, and ORF1ab gene regions. The second targets the viral N and E-gene regions, and the third targets the ORF1ab and E-gene regions.

All PCR testing was conducted at the Hamad Medical Corporation Central Laboratory or Sidra Medicine Laboratory, following standardized protocols.

### Rapid antigen testing

Severe acute respiratory syndrome coronavirus 2 (SARS-CoV-2) antigen tests were performed on nasopharyngeal swabs using one of the following lateral flow antigen tests: Panbio COVID-19 Ag Rapid Test Device (Abbott, USA); SARS-CoV-2 Rapid Antigen Test (Roche, Switzerland); Standard Q COVID-19 Antigen Test (SD Biosensor, Korea); or CareStart COVID-19 Antigen Test (Access Bio, USA). All antigen tests were performed point-of-care according to each manufacturer’s instructions at public or private hospitals and clinics throughout Qatar with prior authorization and training by the Ministry of Public Health (MOPH). Antigen test results were electronically reported to the MOPH in real time using the Antigen Test Management System which is integrated with the national Coronavirus Disease 2019 (COVID-19) database.

### Classification of infections by variant type

Surveillance for SARS-CoV-2 variants in Qatar is based on viral genome sequencing and multiplex RT-qPCR variant screening^1^ of random positive clinical samples,^2-7^ complemented by deep sequencing of wastewater samples.^4,8,9^ Further details on the viral genome sequencing and multiplex RT-qPCR variant screening throughout the SARS-CoV-2 waves in Qatar can be found in previous publications.^2-7,10-16^

## Section S2. COVID-19 severity, criticality, and fatality classification

Classification of COVID-19 case severity (acute-care hospitalizations),^17^ criticality (intensive-care-unit hospitalizations),^17^ and fatality^18^ followed World Health Organization (WHO) guidelines. Assessments were made by trained medical personnel independent of study investigators and using individual chart reviews, as part of a national protocol applied to every hospitalized COVID-19 patient. Each hospitalized COVID-19 patient underwent an infection severity assessment every three days until discharge or death. We classified individuals who progressed to severe, critical, or fatal COVID-19 between the time of the documented infection and the end of the study based on their worst outcome, starting with death,^18^ followed by critical disease,^17^ and then severe disease.^17^

Severe COVID-19 disease was defined per WHO classification as a SARS-CoV-2 infected person with “oxygen saturation of <90% on room air, and/or respiratory rate of >30 breaths/minute in adults and children >5 years old (or ≥60 breaths/minute in children <2 months old or ≥50 breaths/minute in children 2-11 months old or ≥40 breaths/minute in children 1–5 years old), and/or signs of severe respiratory distress (accessory muscle use and inability to complete full sentences, and, in children, very severe chest wall indrawing, grunting, central cyanosis, or presence of any other general danger signs)”.^17^ Detailed WHO criteria for classifying SARS-CoV-2 infection severity can be found in the WHO technical report.^17^

Critical COVID-19 disease was defined per WHO classification as a SARS-CoV-2 infected person with “acute respiratory distress syndrome, sepsis, septic shock, or other conditions that would normally require the provision of life sustaining therapies such as mechanical ventilation (invasive or non-invasive) or vasopressor therapy”.^17^ Detailed WHO criteria for classifying SARS-CoV-2 infection criticality can be found in the WHO technical report.^17^

COVID-19 death was defined per WHO classification as “a death resulting from a clinically compatible illness, in a probable or confirmed COVID-19 case, unless there is a clear alternative cause of death that cannot be related to COVID-19 disease (e.g. trauma). There should be no period of complete recovery from COVID-19 between illness and death. A death due to COVID-19 may not be attributed to another disease (e.g. cancer) and should be counted independently of preexisting conditions that are suspected of triggering a severe course of COVID-19”. Detailed WHO criteria for classifying COVID-19 death can be found in the WHO technical report.^18^

**Figure S1.**
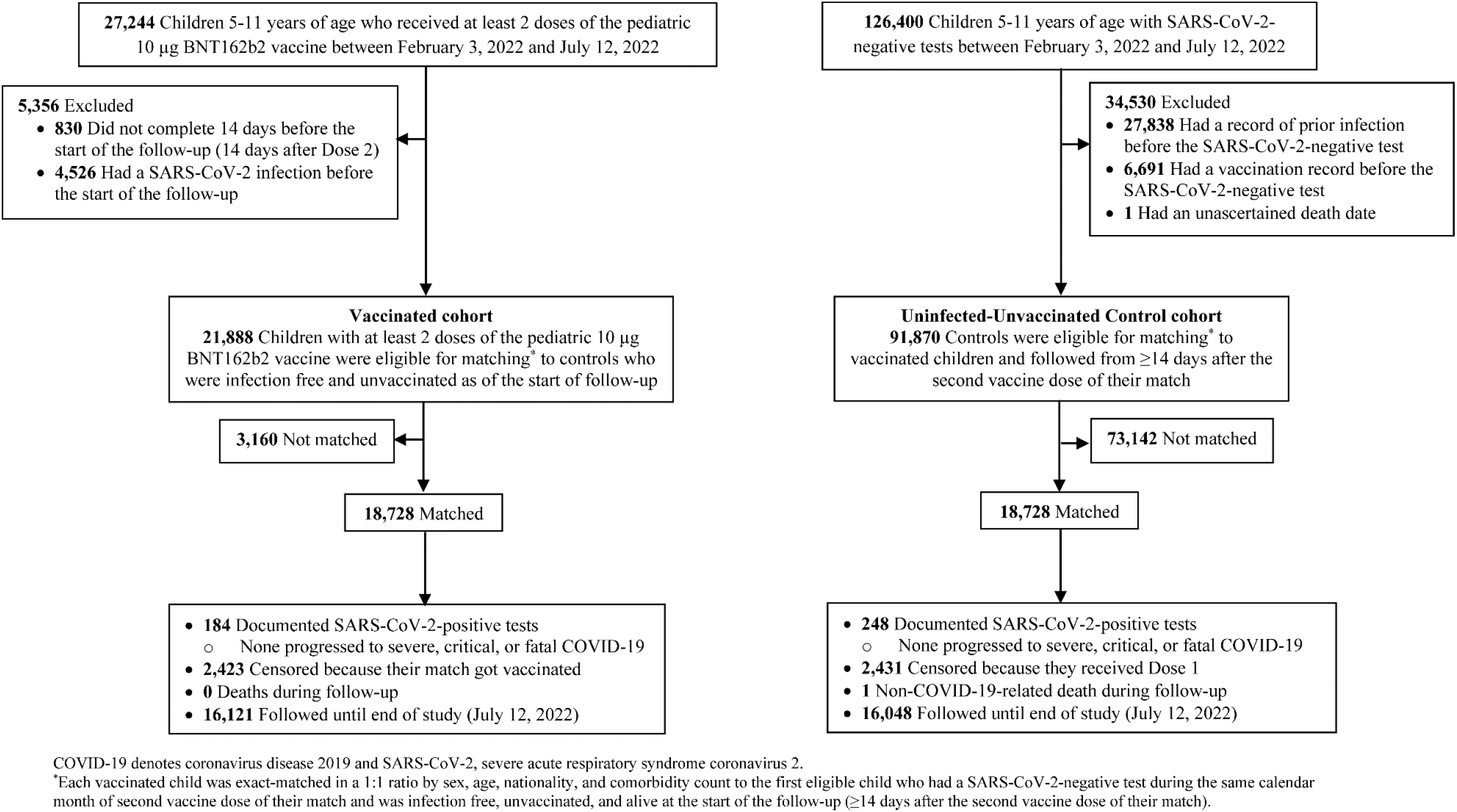
**Cohort selection for investigating effectiveness of the pediatric 10 µg BNT162b2 vaccine among children aged 5-11 years against infection with an Omicron subvariant in Qatar (Children Omicron 10 µg BNT162b2 Study). 6**

**Figure S2.**
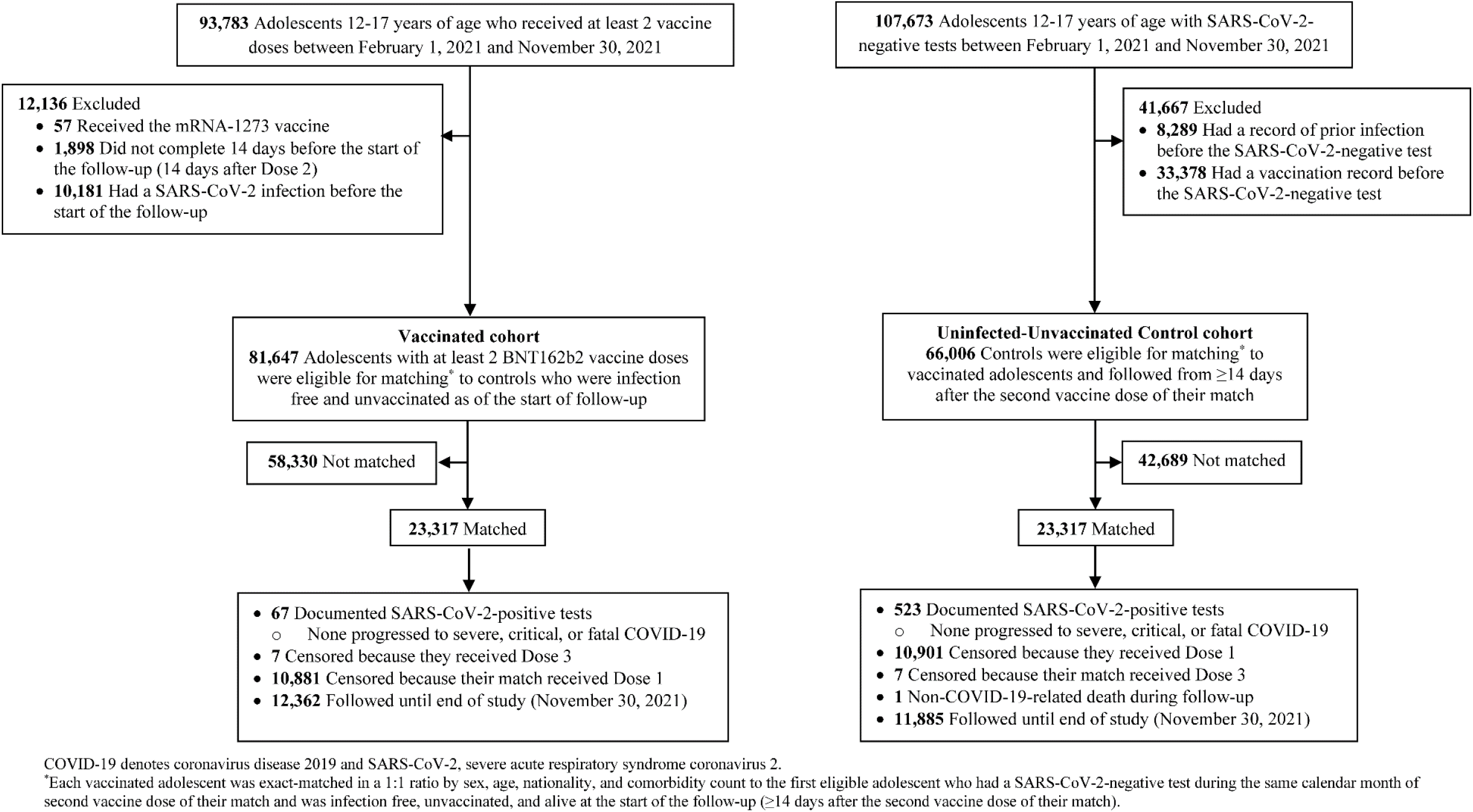
**Cohort selection for investigating effectiveness of the 30 µg BNT162b2 vaccine among adolescents aged 12-17 years against infection with a pre-Omicron variant in Qatar (Adolescents Pre-Omicron 30 µg BNT162b2 Study) 7**

**Figure S3.**
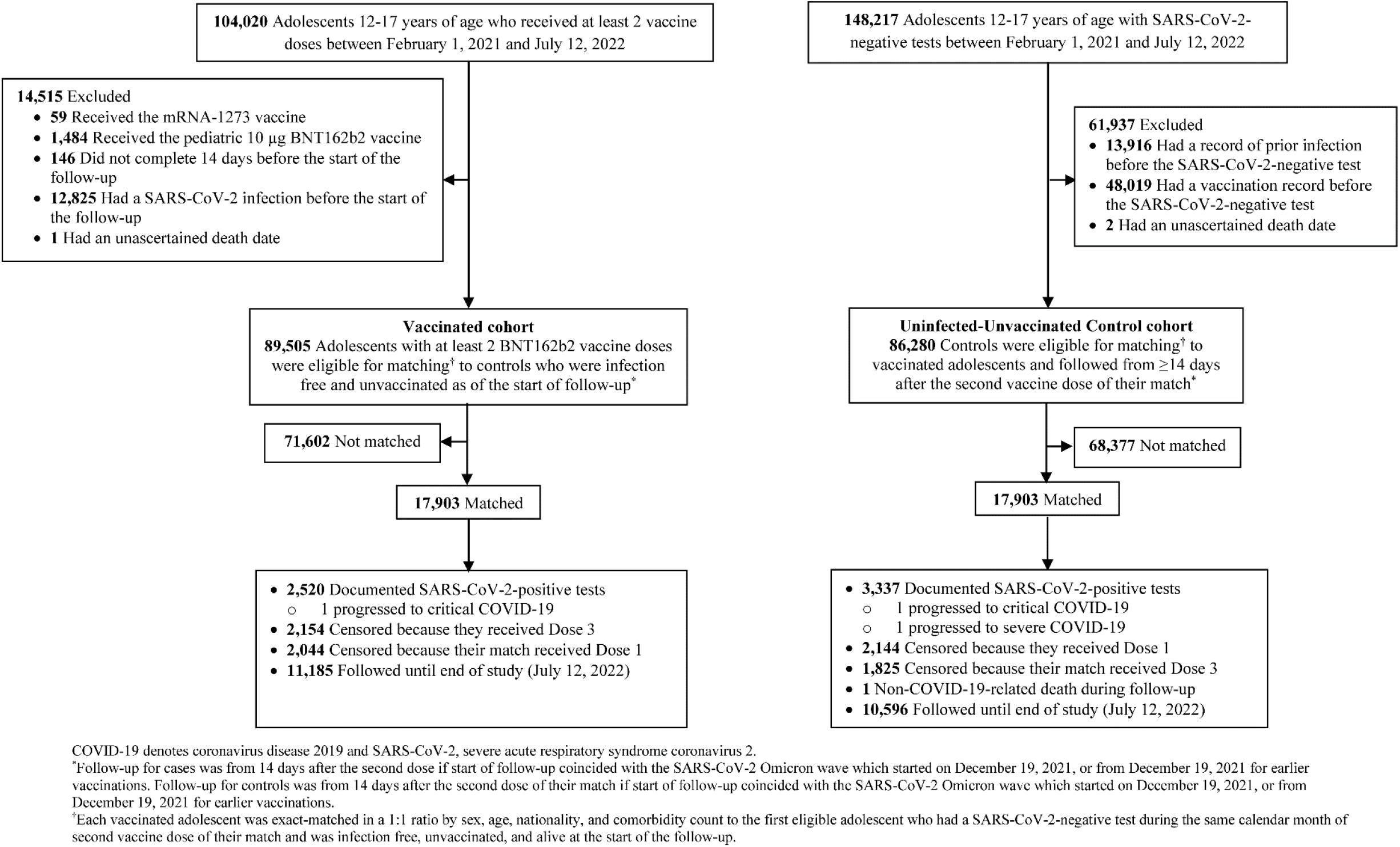
**Cohort selection for investigating effectiveness of the 30 µg BNT162b2 vaccine among adolescents aged 12-17 years against infection with an Omicron subvariant in Qatar (Adolescents Omicron 30 µg BNT162b2 Study). 8**

**Table S1.**
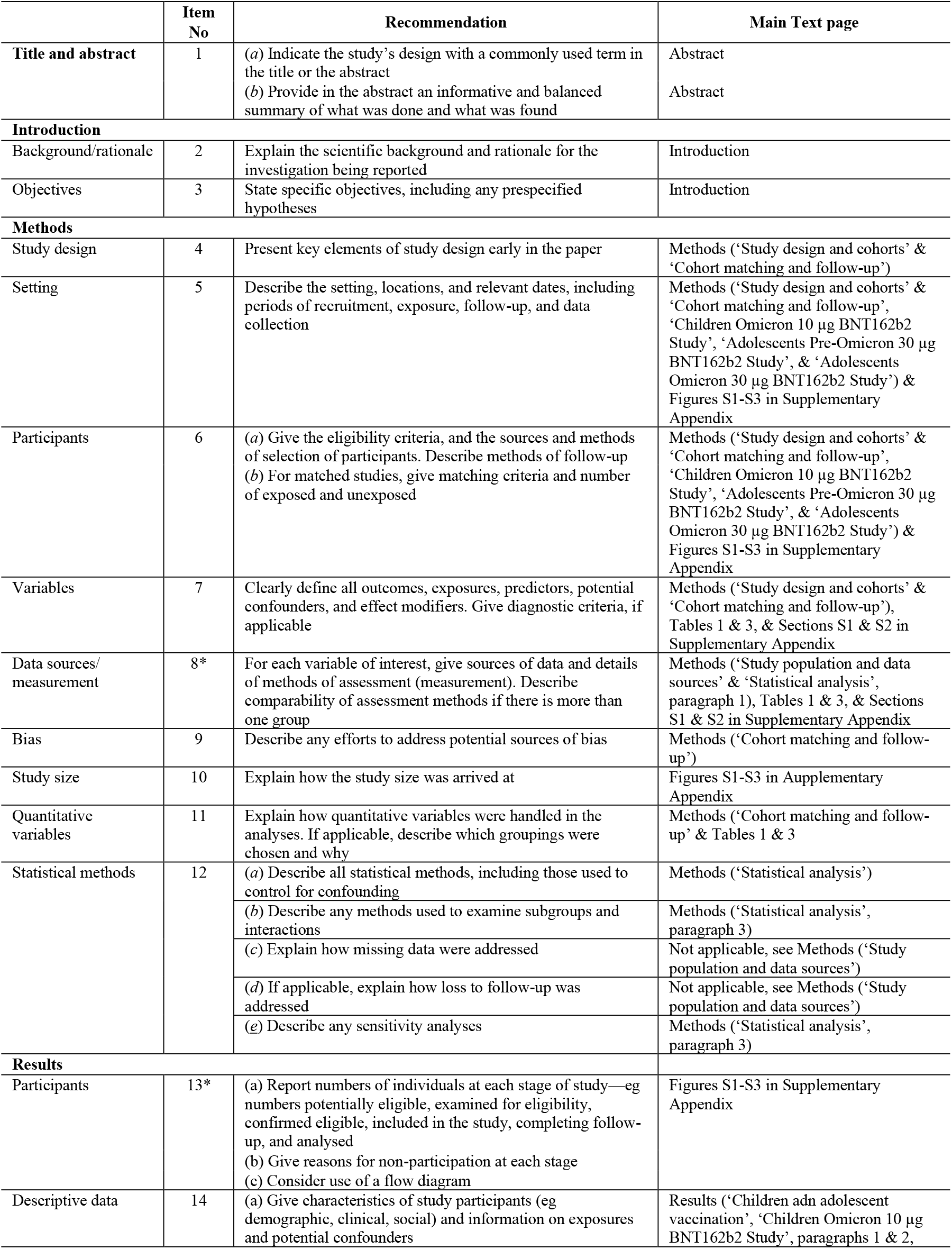
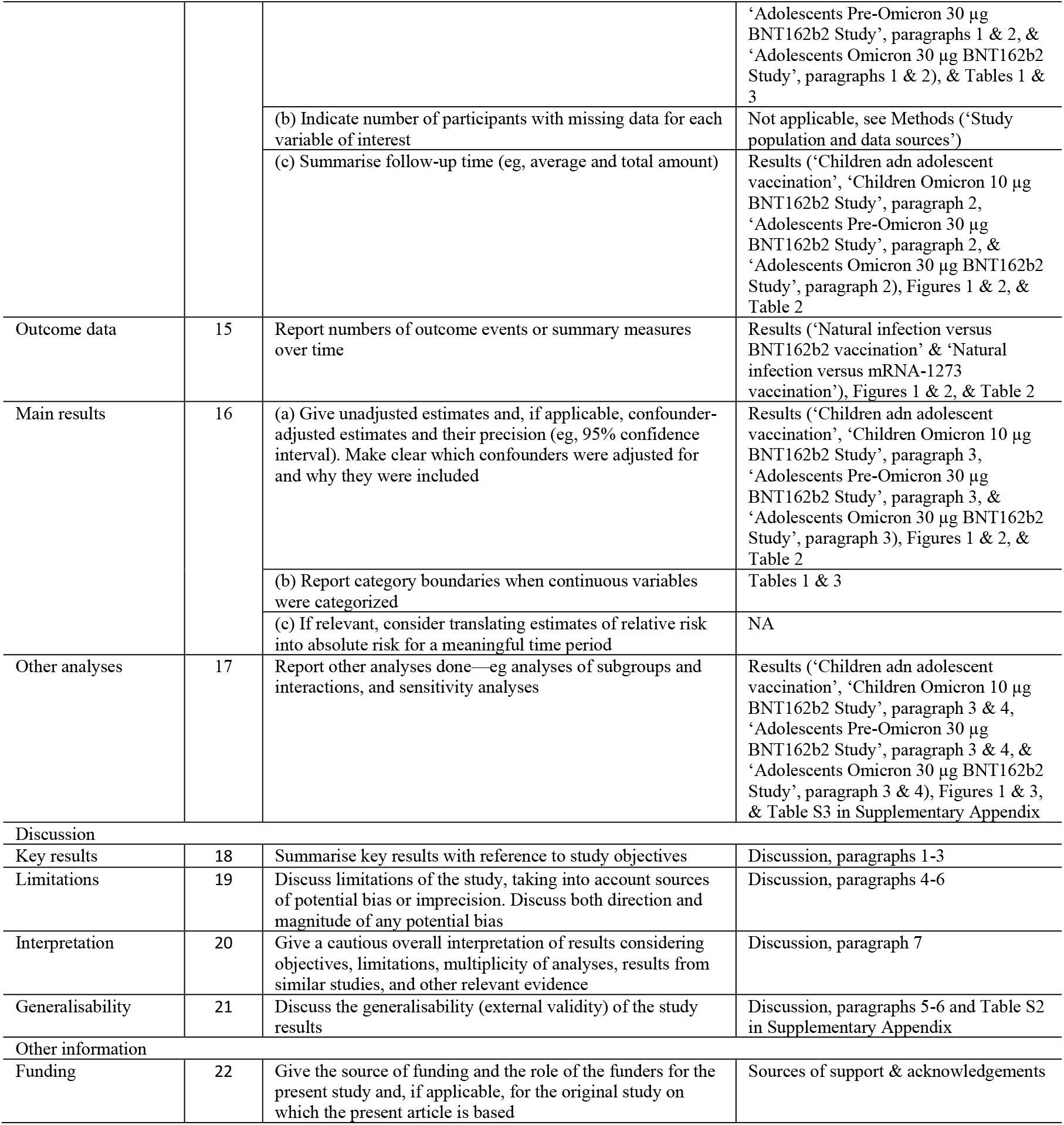
STROBE checklist for cohort studies.

**Table S2.** Representativeness of study participants.

|  |  |
| --- | --- |
| **Category** |  |
| Disease, problem, or condition under investigation | Protection conferred by the two-dose regimens of the pediatric 10 µg BNT162b2 vaccine and of the 30 µg BTN162b2 vaccine among children aged 5-11 years and adolescents aged 12-17 years, respectively, against breakthrough infection with SARS-CoV-2. |
| Special considerations related to<br>Sex and gender | Three national matched, retrospective target-trial cohort studies were conducted to compare incidence of SARS-CoV-2 infection among children or adolescents who completed two-dose vaccination to incidence among those infection-naïve and unvaccinated. Cohorts were exact-matched by sex to control for potential differences in the risk of exposure to SARS-CoV-2 infection by sex. |
| Age | Cohorts were exact-matched by exact age to control for potential differences in the risk of exposure to SARS-CoV-2 infection by age and to account for differences in vaccine effectiveness by age. |
| Race or ethnicity group | Cohorts were exact-matched by nationality to control for potential differences in the risk of exposure to SARS-CoV-2 infection by nationality. Nationality is associated with race and ethnicity in the population of Qatar. |
| Geography | Individual-level data on geography were not available, but Qatar is essentially a city state and infection incidence and vaccination were broadly distributed across the country's neighborhoods/areas. Cohorts were exact-matched by nationality to control for potential differences in the risk of exposure to SARS-CoV-2 infection by nationality. Qatar has unusually diverse demographics in that 89% of the population are international expatriate residents coming from over 150 countries from all world regions. |
| Other considerations | To ensure that all individuals in all cohorts have a record of active presence in Qatar at the same calendar time, individuals who received their second vaccine dose in a specific month were matched to infection-naïve individuals who had a record of a SARS-CoV-2 negative test in that same calendar month. |
| Overall representativeness of this study | The study was based on the total children and adolescents population of Qatar and thus the study population is broadly representative of the diverse, by national background, children and adolescents population of Qatar. While there could be differences in the risk of exposure to SARS-CoV-2 infection by sex, age, nationality, and comorbidity, cohorts were exact-matched by these factors to control for their potential impact on our estimates. |

SARS-CoV-2 denotes severe acute respiratory syndrome coronavirus 2.

**Table S3.** Sensitivity analyses adjusting effectiveness estimates by the testing frequency ratio among the vaccinated and control cohorts.

| Epidemiological measure | Vaccinated cohort* | Control cohort* |
| --- | --- | --- |
| Children Omicron 10 µg BNT162b2 Study |  |  |
| Proportion with a SARS-CoV-2 test during follow-up (%) | 22.7 | 37.5 |
| Testing frequency during follow-up (tests per person) | 0.30 | 0.47 |
| Effectiveness adjusted for ratio in testing frequency (%) | -16.5 (95% CI: -41.0 to 3.7) |  |
| Adolescents Pre-Omicron 30 µg BNT162b2 Study |  |  |
| Proportion with a SARS-CoV-2 test during follow-up (%) | 27.2 | 43.5 |
| Testing frequency during follow-up (tests per person) | 0.39 | 0.64 |
| Effectiveness adjusted for ratio in testing frequency (%) | 79.6 (95% CI: 73.6 to 84.2) |  |
| Adolescents Omicron 30 µg BNT162b2 Study |  |  |
| Proportion with a SARS-CoV-2 test during follow-up (%) | 50.9 | 49.8 |
| Testing frequency during follow-up (tests per person) | 0.89 | 0.84 |
| Effectiveness adjusted for ratio in testing frequency (%) | 33.9 (95% CI: 30.5 to 37.3) |  |

CI denotes confidence interval and SARS-CoV-2 severe acute respiratory syndrome coronavirus 2.
\*The differences in testing frequency between the cohorts were mainly related to different testing guidelines for those vaccinated compared to those unvaccinated, especially in relation to travel.

## References

1. Polack FP, Thomas SJ, Kitchin N, et al. Safety and Efficacy of the BNT162b2 mRNA Covid-19 Vaccine. N Engl J Med 2020;383:2603–15.

2. Walter EB, Talaat KR, Sabharwal C, et al. Evaluation of the BNT162b2 Covid-19 Vaccine in Children 5 to 11 Years of Age. N Engl J Med 2022;386:35–46.

3. Abu-Raddad LJ, Chemaitelly H, Ayoub HH, et al. Characterizing the Qatar advanced-phase SARS-CoV-2 epidemic. Sci Rep 2021;11:6233.

4. Abu-Raddad LJ, Chemaitelly H, Ayoub HH, et al. Introduction and expansion of the SARS-CoV-2 B.1.1.7 variant and reinfections in Qatar: A nationally representative cohort study. PLoS Med 2021;18:e1003879.

5. Chemaitelly H, Bertollini R, Abu-Raddad LJ, National Study Group for Covid Epidemiology. Efficacy of Natural Immunity against SARS-CoV-2 Reinfection with the Beta Variant. N Engl J Med 2021;385:2585–6.

6. Altarawneh HN, Chemaitelly H, Ayoub HH, et al. Effects of Previous Infection and Vaccination on Symptomatic Omicron Infections. N Engl J Med 2022.

7. Altarawneh HN, Chemaitelly H, Ayoub HH, et al. Protection of SARS-CoV-2 natural infection against reinfection with the Omicron BA.4 or BA.5 subvariants. medRxiv 2022:2022.07.11.22277448.

8. Tang P, Hasan MR, Chemaitelly H, et al. BNT162b2 and mRNA-1273 COVID-19 vaccine effectiveness against the SARS-CoV-2 Delta variant in Qatar. Nat Med 2021;27:2136–43.

9. Chemaitelly H, Tang P, Hasan MR, et al. Waning of BNT162b2 Vaccine Protection against SARS-CoV-2 Infection in Qatar. N Engl J Med 2021;385:e83.

10. Abu-Raddad LJ, Chemaitelly H, Ayoub HH, et al. Effect of mRNA Vaccine Boosters against SARS-CoV-2 Omicron Infection in Qatar. N Engl J Med 2022;386:1804–16.

11. Hernan MA, Robins JM. Using Big Data to Emulate a Target Trial When a Randomized Trial Is Not Available. Am J Epidemiol 2016;183:758–64.

12. World Health Organization. COVID-19 clinical management: living guidance. Available from: https://www.who.int/publications/i/item/WHO-2019-nCoV-clinical-2021-1. Accessed on: May 15, 2021. 2021.

13. World Health Organization. International guidelines for certification and classification (coding) of COVID-19 as cause of death. Available from: https://www.who.int/classifications/icd/Guidelines_Cause_of_Death_COVID-19-20200420-EN.pdf?ua=1. Document Number: WHO/HQ/DDI/DNA/CAT. Accessed on May 15, 2021. 2020.

14. Ayoub HH, Chemaitelly H, Seedat S, et al. Mathematical modeling of the SARS-CoV-2 epidemic in Qatar and its impact on the national response to COVID-19. J Glob Health 2021;11:05005.

15. Coyle PV, Chemaitelly H, Ben Hadj Kacem MA, et al. SARS-CoV-2 seroprevalence in the urban population of Qatar: An analysis of antibody testing on a sample of 112,941 individuals. iScience 2021;24:102646.

16. Al-Thani MH, Farag E, Bertollini R, et al. SARS-CoV-2 Infection Is at Herd Immunity in the Majority Segment of the Population of Qatar. Open Forum Infect Dis 2021;8:ofab221.

17. Jeremijenko A, Chemaitelly H, Ayoub HH, et al. Herd Immunity against Severe Acute Respiratory Syndrome Coronavirus 2 Infection in 10 Communities, Qatar. Emerg Infect Dis 2021;27:1343–52.

18. Abu-Raddad LJ, Chemaitelly H, Yassine HM, et al. Pfizer-BioNTech mRNA BNT162b2 Covid-19 vaccine protection against variants of concern after one versus two doses. J Travel Med 2021;28.

19. Chemaitelly H, Yassine HM, Benslimane FM, et al. mRNA-1273 COVID-19 vaccine effectiveness against the B.1.1.7 and B.1.351 variants and severe COVID-19 disease in Qatar. Nat Med 2021;27:1614–21.

20. Abu-Raddad LJ, Chemaitelly H, Bertollini R, National Study Group for Covid Vaccination. Waning mRNA-1273 Vaccine Effectiveness against SARS-CoV-2 Infection in Qatar. N Engl J Med 2022.

21. Abu-Raddad LJ, Chemaitelly H, Bertollini R, National Study Group for Covid Vaccination. Effectiveness of mRNA-1273 and BNT162b2 Vaccines in Qatar. N Engl J Med 2022.

22. Barda N, Dagan N, Cohen C, et al. Effectiveness of a third dose of the BNT162b2 mRNA COVID-19 vaccine for preventing severe outcomes in Israel: an observational study. Lancet 2021;398:2093–100.

23. Altarawneh HN, Chemaitelly H, Hasan MR, et al. Protection against the Omicron Variant from Previous SARS-CoV-2 Infection. N Engl J Med 2022.

24. Austin PC. Using the Standardized Difference to Compare the Prevalence of a Binary Variable Between Two Groups in Observational Research. Communications in Statistics - Simulation and Computation 2009;38:1228–34.

25. Kaplan EL, Meier P. Nonparametric Estimation from Incomplete Observations. J Am Stat Assoc 1958;53:457–81.

26. Hasan MR, Kalikiri MKR, Mirza F, et al. Real-Time SARS-CoV-2 Genotyping by High-Throughput Multiplex PCR Reveals the Epidemiology of the Variants of Concern in Qatar. Int J Infect Dis 2021;112:52–4.

27. Benslimane FM, Al Khatib HA, Al-Jamal O, et al. One Year of SARS-CoV-2: Genomic Characterization of COVID-19 Outbreak in Qatar. Front Cell Infect Microbiol 2021;11:768883.

28. Chemaitelly H, Ayoub HH, AlMukdad S, et al. Duration of mRNA vaccine protection against SARS-CoV-2 Omicron BA.1 and BA.2 subvariants in Qatar. Nat Commun 2022;13:3082.

29. Fowlkes AL, Yoon SK, Lutrick K, et al. Effectiveness of 2-Dose BNT162b2 (Pfizer BioNTech) mRNA Vaccine in Preventing SARS-CoV-2 Infection Among Children Aged 5-11 Years and Adolescents Aged 12-15 Years - PROTECT Cohort, July 2021-February 2022. MMWR Morb Mortal Wkly Rep 2022;71:422–8.

30. Cohen-Stavi CJ, Magen O, Barda N, et al. BNT162b2 Vaccine Effectiveness against Omicron in Children 5 to 11 Years of Age. N Engl J Med 2022;387:227–36.

31. Veneti L, Berild JD, Watle SV, et al. Vaccine effectiveness with BNT162b2 (Comirnaty, Pfizer-BioNTech) vaccine against reported SARS-CoV-2 Delta and Omicron infection among adolescents, Norway, August 2021 to January 2022. medRxiv 2022:2022.03.24.22272854.

32. Kildegaard H, Lund LC, Hojlund M, Stensballe LG, Pottegard A. Risk of adverse events after covid-19 in Danish children and adolescents and effectiveness of BNT162b2 in adolescents: cohort study. BMJ 2022;377:e068898.

33. Sacco C, Del Manso M, Mateo-Urdiales A, et al. Effectiveness of BNT162b2 vaccine against SARS-CoV-2 infection and severe COVID-19 in children aged 5-11 years in Italy: a retrospective analysis of January-April, 2022. Lancet 2022;400:97–103.

34. Seedat S, Chemaitelly H, Ayoub HH, et al. SARS-CoV-2 infection hospitalization, severity, criticality, and fatality rates in Qatar. Sci Rep 2021;11:18182.

35. Butt AA, Dargham SR, Loka S, et al. COVID-19 Disease Severity in Children Infected with the Omicron Variant. Clin Infect Dis 2022.

36. Butt AA, Dargham SR, Tang P, et al. COVID-19 disease severity in persons infected with the Omicron variant compared with the Delta variant in Qatar. J Glob Health 2022;12:05032.

37. Ray GT, Lewis N, Klein NP, Daley MF, Lipsitch M, Fireman B. Depletion-of-susceptibles Bias in Analyses of Intra-season Waning of Influenza Vaccine Effectiveness. Clin Infect Dis 2020;70:1484–6.

## References

1. Multiplexed RT-qPCR to screen for SARS-COV-2 B.1.1.7, B.1.351, and P.1 variants of concern V.3. dx.doi.org/10.17504/protocols.io.br9vm966. 2021. (Accessed June 6, 2021, at https://www.protocols.io/view/multiplexed-rt-qpcr-to-screen-for-sars-cov-2-b-1-1-br9vm966.)

2. Abu-Raddad LJ, Chemaitelly H, Butt AA, National Study Group for Covid Vaccination. Effectiveness of the BNT162b2 Covid-19 Vaccine against the B.1.1.7 and B.1.351 Variants. N Engl J Med 2021;385:187–9.

3. Chemaitelly H, Yassine HM, Benslimane FM, et al. mRNA-1273 COVID-19 vaccine effectiveness against the B.1.1.7 and B.1.351 variants and severe COVID-19 disease in Qatar. Nat Med 2021;27:1614–21.

4. Qatar viral genome sequencing data. Data on randomly collected samples. https://www.gisaid.org/phylodynamics/global/nextstrain/. 2021. at https://www.gisaid.org/phylodynamics/global/nextstrain/.)

5. Benslimane FM, Al Khatib HA, Al-Jamal O, et al. One Year of SARS-CoV-2: Genomic Characterization of COVID-19 Outbreak in Qatar. Front Cell Infect Microbiol 2021;11:768883.

6. Hasan MR, Kalikiri MKR, Mirza F, et al. Real-Time SARS-CoV-2 Genotyping by High-Throughput Multiplex PCR Reveals the Epidemiology of the Variants of Concern in Qatar. Int J Infect Dis 2021;112:52–4.

7. Chemaitelly H, Tang P, Hasan MR, et al. Waning of BNT162b2 Vaccine Protection against SARS-CoV-2 Infection in Qatar. N Engl J Med 2021;385:e83.

8. Saththasivam J, El-Malah SS, Gomez TA, et al. COVID-19 (SARS-CoV-2) outbreak monitoring using wastewater-based epidemiology in Qatar. Sci Total Environ 2021;774:145608.

9. El-Malah SS, Saththasivam J, Jabbar KA, et al. Application of human RNase P normalization for the realistic estimation of SARS-CoV-2 viral load in wastewater: A perspective from Qatar wastewater surveillance. Environ Technol Innov 2022;27:102775.

11. Tang P, Hasan MR, Chemaitelly H, et al. BNT162b2 and mRNA-1273 COVID-19 vaccine effectiveness against the SARS-CoV-2 Delta variant in Qatar. Nat Med 2021;27:2136–43.

12. Altarawneh HN, Chemaitelly H, Ayoub HH, et al. Effects of Previous Infection and Vaccination on Symptomatic Omicron Infections. N Engl J Med 2022.

13. Altarawneh HN, Chemaitelly H, Hasan MR, et al. Protection against the Omicron Variant from Previous SARS-CoV-2 Infection. N Engl J Med 2022.

14. Chemaitelly H, Ayoub HH, AlMukdad S, et al. Duration of mRNA vaccine protection against SARS-CoV-2 Omicron BA.1 and BA.2 subvariants in Qatar. Nat Commun 2022;13:3082.

15. Qassim SH, Chemaitelly H, Ayoub HH, et al. Effects of BA.1/BA.2 subvariant, vaccination, and prior infection on infectiousness of SARS-CoV-2 omicron infections. Journal of Travel Medicine 2022.

16. Altarawneh HN, Chemaitelly H, Ayoub HH, et al. Protection of SARS-CoV-2 natural infection against reinfection with the Omicron BA.4 or BA.5 subvariants. medRxiv 2022:2022.07.11.22277448.

17. World Health Organization. COVID-19 clinical management: living guidance. Available from: https://www.who.int/publications/i/item/WHO-2019-nCoV-clinical-2021-1. Accessed on: May 15, 2021. 2021.

18. World Health Organization. International guidelines for certification and classification (coding) of COVID-19 as cause of death. Available from: https://www.who.int/classifications/icd/Guidelines_Cause_of_Death_COVID-19-20200420-EN.pdf?ua=1. Document Number: WHO/HQ/DDI/DNA/CAT. Accessed on May 15, 2021. 2020.

